# After the Haiti 2021 Earthquake: Triage and treatment data for a cohort of patients transported for higher level of care

**DOI:** 10.1101/2021.11.13.21266308

**Authors:** Kelly Edwards, David Ellis, Cory Oaks, SGM Clayton W. Mann, Vincent DeGennaro

## Abstract

**Intro:** On August 14, a magnitude 7.2 earthquake occurred in the South of Haiti, killing over 2,200 people and leaving at least 12,000 injured. On August 17th, a joint task force coordinated the triage, treatment, and transport for patients arriving at the Port-au-Prince airport from the South.

**Methods:** From August 17th-27th, approximately 243 patients were encountered at the Port-au-Prince airport. For the first three days of operations, written notes, reports, and various chat logs from WhatsApp were used to compile this data.

**Results:** 243 patents were encountered at the airport Triage and Treatment from August 17 to August 27. Orthopedic injuries were the most common presenting injury with 20% of patients having a pelvis fracture (the most prevalent injury). Of the 174 with known transfer destinations, 118 (68%) were transported to one of three tertiary care hospitals, with 99 going via ground ambulance to the two in Port au Prince, and 19 going via HAA helicopter to Mirebalais. Once treatment operations were initiated, 106 patients received some form of treatment at the airport while awaiting transport.

**Discussion:** Interagency coordination was the key to rapid scale up. To address the most prevalent patient issues, a treatment bundle was conceptualized a few days into operations that consisted of IV fluids, analgesia, IV antibiotics, wound debridement and cleaning, and tetanus prophylaxis.

## Intro

On August 14, a magnitude 7.2 earthquake occurred in the South of Haiti, killing over 2,200 people and leaving at least 12,000 injured. Due to damaged roads and bridges--made worse by a Tropical Storm-- and gang violence along the route, terrestrial ambulance evacuation of critically-ill patients was not possible. However, the volume of patients coming out of the South by air soon overwhelmed the Port au Prince airport, and patients often sat for hours awaiting transport and usually not getting treatment at the receiving hospital for hours more upon arrival. On August 17^th^, Haiti Air Ambulance (HAA) partnered with Haiti’s National Center of Ambulances (Centre Ambulancier National (CAN)), Haitian Red Cross, US Military through Joint Task Force Haiti, and Hero Client Rescue to coordinate the triage, treatment, and transport for patients arriving at the Port-au-Prince airport from the South. This paper seeks to quantify the rescue efforts to shed light on the post-disaster chaos and to help inform future efforts in similar environments.

## Methods

From August 17^th^-27th, approximately 243 patients were encountered at the Port-au-Prince airport. For the first three days of operations, written notes, reports, and various chat logs from WhatsApp were used to compile this data. From August 20^th^ on, all data was archived and compiled in private, dedicated WhatsApp groups specific for this effort. Communications Specialists from HAA received basic patient information from HAA staff at the airport and arranged acceptance for these patients at appropriate hospitals. All patient data from the airport was included and no patients were intentionally excluded.

## Results

243 patents were encountered at the airport Triage and Treatment from August 17 to August 27. There were 165 adults (68%), 31 (13%) over 65 years old, and 49 (20%) were under 13 years old. Roughly 30% of the patients came from Jeremie and Les Cayes each, the two largest cities in the South. Orthopedic injuries were the most common presenting injury with 20% of patients having a pelvis fracture (the most prevalent injury). 178 of the 243 patients had more than one significant injury, while only 39 (16%) were considered to have only minor injuries. Of the 174 with known transfer destinations, 118 (68%) were transported to one of three tertiary care hospitals, with 99 going via ground ambulance to the two in Port au Prince, and 19 going via HAA helicopter to Mirebalais, 32 miles away. Seventeen patients (10%) were discharged from the airport and released to a nearby shelter where they could await transport back home. Once treatment operations were initiated, 106 patients received some form of treatment at the airport while awaiting transport. This included pain medication, IV fluids, antibiotics, simple wound care, and tetanus prophylaxis. Although treatment was limited, it was crucial for many of these patients who had received little to no care prior to arriving at the airport with injuries that were many days old.

## Discussion

Interagency coordination was the key to rapid scale up. The US Military leveraged their larger helicopters to transport multiple patients and families per trip while HAA utilized its single patient helicopter to transport patients from the airport triage and treatment to more remote receiving hospitals. CAN, Hero Client Rescue, and the Haitian Red Cross provided ground transport services to local receiving hospitals. HAA provided the local expertise on facility capability and treatment options, HERO Client Rescue and ancillary personnel from the tertiary care hospitals drove the triage/treatment/transport decisions with additional staff (medical and non-medical), resources, and advice supplied by the US Military. The rescue efforts were initially hampered by the arrival of a Tropical Storm system on August 16 that resulted in heavy rainfall, slowing air transport, and flooding that caused additional damage to roads and bridges.

To address the most prevalent patient issues, a treatment bundle was conceptualized a few days into operations that consisted of IV fluids, analgesia, IV antibiotics, wound debridement and cleaning, and tetanus prophylaxis. Point-of-care ultrasound was used nine times, with six instances confirming that a patient had free fluid in their abdomen consistent with internal bleeding, allowing proper triaging to accepting hospitals.

This report is limited by less than systematic record keeping for the first three days. The sample is biased towards those ill enough to require transport out of the South by air. This experience, along with Hurricane Matthew in 2016 that also struck the South of Haiti, further validates the need for and clinical utility of helicopter emergency medical services in low-income settings where road infrastructure and terrestrial ambulance services are lacking. The experience is generalizable to low-income countries with poor road and emergency services infrastructure in a post-disaster setting.

**Table 1:** Injuries and Treatment

| **<u>Injury</u>** | **<u>% of total (N)</u>** |
| --- | --- |
| Pelvis Fracture | 20% (49) |
| Minor Injuries | 16% (39) |
| Head Injury | 11.9% (29) |
| Closed Femur Fracture | 11.5% (28) |
| Other | 9.1% (22) |
| Closed Arm/Wrist Fracture | 7.8% (19) |
| Closed Tib-Fib Fracture | 7.8% (19) |
| Open/Infected Wounds | 7% (17) |
| Abdominal Trauma | 6.6% (16) |
| Open Tib-Fib | 6.6% (16) |
| Altered LOC | 5.3% (13) |
| Open Femur Fracture | 3.3% (8) |
| Closed Ankle/Foot Fracture | 2.9% (7) |
| Open Arm/Wrist Fracture | 2.9% (7) |
| Spinal Injury | 2.9% (7) |
| Tetanus | 2.9% (7) |
| Clavicle Fracture | 2.1% (5) |
| Rib Fracture | 2.1% (5) |
| Shoulder Dislocation | 2.1% (5) |
| Back Pain | 1.2% (3) |
| Facial/Jaw Fracture | 1.2% (3) |
| Lacerations/Abrasion | 1.2% (3) |
| Skull Fracture | 1.2% (3) |
| Burns | 0.8% (2) |
| Crush Syndrome | 0.8% (2) |

| **<u>Treatment</u>** | **<u>% of total (N)</u>** |
| --- | --- |
| Fluids, Analgesia only | 30.1% (32) |
| Treatment Bundle (Fluids, antibiotics, tetanus, analgesia) | 28.3% (30) |
| Analgesia | 14.1% (15) |
| Oral Amoxicillin Only | 12.3% (13) |
| Fluids Only | 9.4% (10) |
| Fluids, Analgesia, Antibiotics | 1.9% (2) |
| Fluids, Antibiotics | 1.9% (2) |

| **<u>Item/Procedure</u>** | **<u>Number</u>** |
| --- | --- |
| Doses of IV Analgesia | 79 |
| Liters of IV Fluid Administered | 76 |
| Vials of Ceftriaxone Used | 68 |
| Splints Used | 40 |
| Tetanus Antitoxin | 31 |
| Wound Debridement | 31 |
| Ultrasound Performed | 9 |

